# Knockdown resistance (kdr) Associated organochlorine Resistance in human head lice: Systematic review and meta-analysis

**DOI:** 10.1101/2022.10.02.22280631

**Authors:** Ebrahim Abbasi, Zahra Nasiri, Jalal Mohammadi, Zahra Yazdani, Shokrollah Mohseni

## Abstract

**Introduction:** Pediculosis is one of the vector-borne diseases that has spread in most regions of the world and has affected many populations. In previous decades, organochlorine poisons were used to treat it. But resistance to treatment against this group of insecticides affected its control. Based on this, the present study was conducted to investigate the prevalence of Knockdown Resistance in human head lice against organochlorine insecticides in the world in the form of a systematic review and meta-analysis.

**Methods:** To determine the prevalence of Knockdown Resistance against organochlorine insecticides, all English and non-English articles (at least with English titles and abstracts) were published worldwide without a time limit until the end of May 2022 and were extracted and analyzed. Statistical analysis of the data was done using statistical tests of fixed and random effects model in the meta-analysis, Cochrane, meta-regression, and I2 index.

**Results:** 8 articles with a sample size of 7249 head lice were included in the meta-analysis process. The prevalence of knockdown resistance was estimated at 65.3%. Also, the prevalence of homozygote resistance was 71.9% and heterozygote resistance was 28.1%.

**Conclusion:** Based on the findings, more than half of the lice were resistant to organochlorine insecticides. As a result, due to the high prevalence of resistance, it is recommended to determine the resistance against them before using this group of insecticides to treat pediculosis and then adopting the appropriate treatment.

## Introduction

Head lice are one of the most common infections around the world, which infects mostly school-age children and adults through head-to-head transmission, and if untreated, it can widely affect most people in communities, which leads to the imposition of costs. It greatly affects the health, medical systems, and families .(2, 1)Various factors affect the increase in the prevalence of this contamination in the world, including being in large and contaminated populations such as schools or families, lack of awareness and compliance with health principles, the number and duration of contact with infected people, and finally inappropriate treatment. and pointed out incomplete contamination .(4, 3)

One of the most important approaches to controlling pediculosis is treating infected people. Because with the treatment of these people, the source of contamination is eliminated and in addition to removing the contamination in the individual, it is also prevented from spreading to other people, and ultimately the spread of the disease is reduced. Therefore, complete and effective treatment of pediculosis is the most important measure to prevent its spread. The most effective treatment method is the use of safe insecticides, including organophosphorus, organochlorine, carbamates, and pyrethroids, which kill the lice by affecting the nervous system or removing their protective covering. These insecticides are widely used in the world in the form of shampoos or tablets with the brand names Permethrin 1%, Malatinon 0.5%, Lindane 1%, and Crotamiton 10% .(7-5).But recently, due to excessive use and incomplete treatment, lice have become resistant to these insecticides .(8) One of the most important mechanisms of the effect of insecticides on lice to eliminate them is the nervous mechanism. So that these insecticides connect to the nervous system by acting on sodium channels sensitive to voltage and cause these channels to remain open for a long time. Finally, due to the long-term absence of these channels, sodium enters uncontrollably and leads to nerve depolarization and hyperpolarization, which ultimately leads to muscle paralysis (knockdown) and the death of the louse. .(10, 9)In the past decades, more than 50 mutations in the sodium channel have been identified individually or in combination, which has been associated with the development of resistance in lice. The most important resistance associated with these mutations is Knockdown resistance (kdr). KDR reduces nerve sensitivity and dysfunction of sodium channels to insecticides. This decrease in sensitivity is caused by point mutations in the genetic composition of insects. Such mutational resistance is characterized by the presence of the kdr allele in the insect genome-11).(13

Insecticides with organochlorine composition were used as an old form of insecticides to deal with insects. But the commercial forms of DDT, Dieldrin, or Chlordcon are not used due to other biological issues, and lindane is used to fight lice. Organochlorine insecticides target the nervous system of insects and sodium channels, and by keeping them open for a long time, they increase their action potential and lead to excessive excitability, muscle paralysis, and death. to be Also, by blocking the GABA-A receptor, this insecticide leads to convulsions in the chloride channels. Therefore, the creation of resistance caused by kdr reduces the effect of these insecticides and the treatment of contamination faces a problem .(16-14) Awareness of the prevalence of human head louse resistance to insecticides is necessary to decide whether to use an appropriate insecticide individually or in combination to treat, control, and reduce the prevalence of infestation. Based on this, the present study was conducted to determine the prevalence of kdr resistance against organochlorine insecticides in human head lice in the form of a systematic review and meta-analysis in the world.

## Analyzation method

This systematic review and meta-analysis study was conducted based on the Preferred Reporting Items for Systematic Reviews and Meta-Analyses (PRISMA) guidelines on the prevalence of kdr in human head lice against organochlorine insecticides worldwide. .(17)The protocol of this study has been registered in the International Prospective Register of Systematic Reviews (PROSPERO) with the code CRD42021231602. Based on this, searching for articles in scientific databases, selecting articles, evaluating the quality of articles, and extracting data were done by two researchers independently.

### Search articles

Search for articles in international scientific databases Web of Science, PubMed, Proquest, Bioone.org, Embase, and Scopus without time limit until the end of May 2022 using keywords Organochlorine, DDT, Dichlorodiphenyl Dichloroethylene, ethane, chlorophenyl, dichloro, ethyl phenyl, Linden, Aldrin, andrin, diendrin, heptachlor, endosulfan, benzene, chlorobenzene, trichloroethylidene, Knockdown resistance, KDR, insecticide, Insecticide Resistance, Pediculicide Resistance, Resistance Mutations, Head Lice, Head Louse, Pediculosis. which were extracted from medical subject headings (Mesh) and were searched in the title, abstract and full text of the articles individually and in combination using OR and AND operators. The search syntax is available in scientific databases.

### Entry and exit criteria

Observational studies, and English-language articles, which investigated resistance to organochlorine toxins in lice, evaluated KDR resistance and investigated its prevalence, were included in the study. Interventional studies, clinical trials, qualitative studies, reviews, letters to the editor, case reports and case series, studies with low quality, and studies related to other insecticides were excluded from the study.

### Quality assessment of articles

The risk of bias was assessed by two independent investigators using the Hoy and JBI instruments designed for prevalence studies. All studies were divided into three categories (low risk–high risk and unknown risk of bias). Any disagreement between researchers was resolved by consensus (18).

### Extracting the Data

To extract the required information, the title and abstract of the articles were independently examined by two researchers based on the inclusion and exclusion criteria of the articles. Then the full text of the selected articles was examined. In case the articles were rejected by the researcher, the reason was mentioned, and in case of disagreement between them, the article was referred to by a third person. To extract data from the checklist, which includes the first author’s characteristics, the article’s time of publication, the study’s location, the prevalence of kdr resistance, and the ratio of types of kdr resistance, was done.

### Selection of studies

At first, references of 14536 articles searched in scientific databases were entered into Endnote software. Then repetition was done and the title and abstract of the articles were examined. Based on this, 8430 articles were excluded due to repetition and 5786 articles due to non-relevance. Then, the text of the reviewed articles and the number of 311 articles were removed due to the lack of investigation of prevalence, lack of investigation of kdr resistance, or lack of investigation of resistance to organochlorine toxins, and 8 articles entered the systematic review and meta-analysis process (Figure 1).

**Figure 1.**
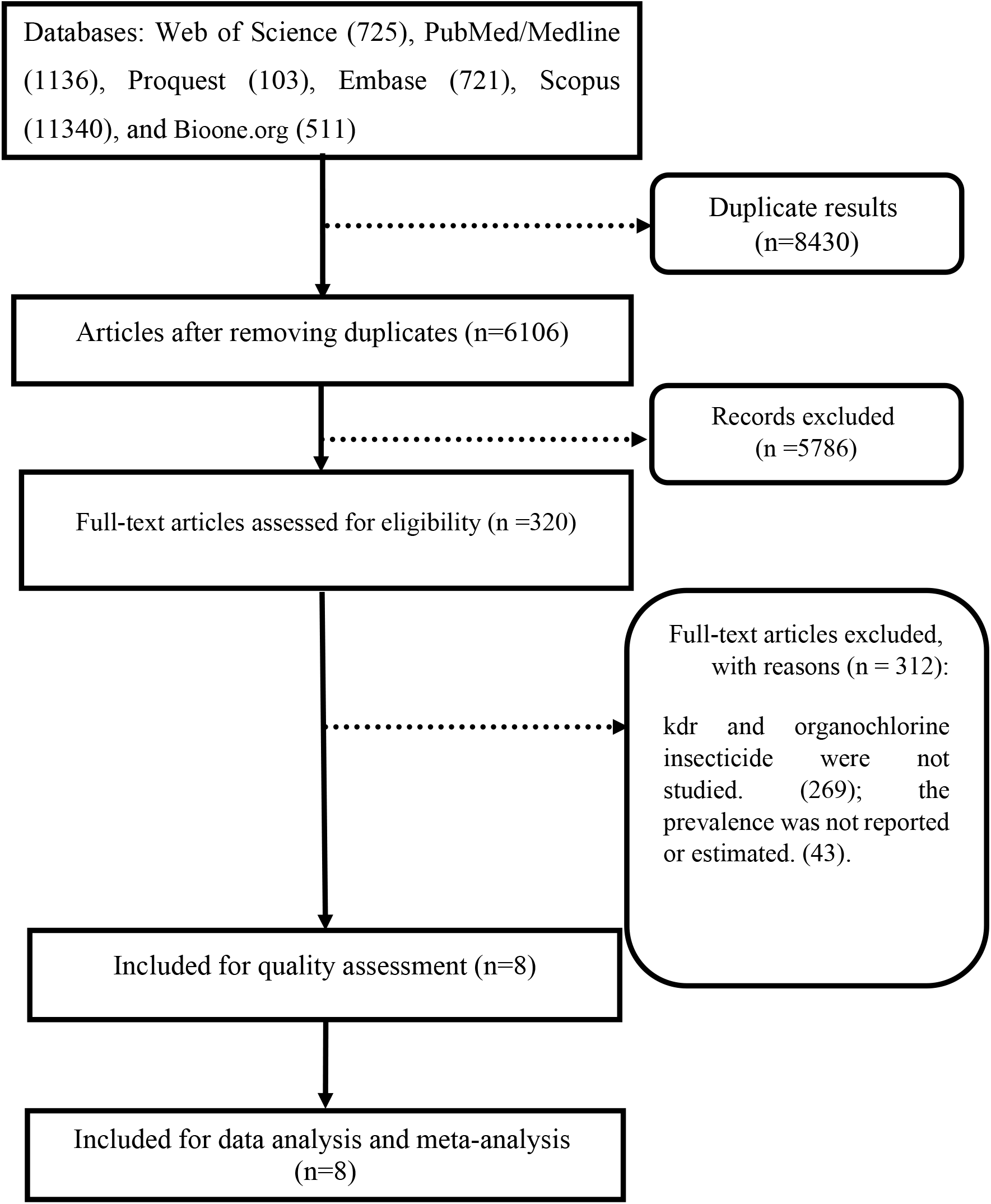
The PRISMA flow diagram

### Statistical analysis of data

The random effects model was used to estimate the prevalence if there were heterogeneous between studies and the fixed effects model was used in the meta-analysis if the studies were homogeneous. The heterogeneity of studies was assessed using Cochrane Q statistics and the I2 test. Accordingly, I2 index was divided into three categories <25% (low heterogeneity), 25-75% (moderate heterogeneity in the study) and> 75% (high heterogeneity). Funnel plot and Egger test were used to examine the publication bias and metaregression was used to investigate the relationship between the year of study and sample size with prevalence. Statistical analysis of data was performed using STATA ver14 software.

## Results

Eight studies with a volume of 7249 head lice that were conducted between 2003 and 2014 were included in the meta-analysis process. The specifications of the reviewed articles are presented in Table 1.

**Table 1.** characteristic of studies included in the systematic review and meta-analysis

| Authors | Year<br>of<br>study | Place of<br>study | Sample<br>size | Prevalence<br>kdr<sup>#</sup> (%) | Prevalence<br>RR<sup>*</sup> (%) | Prevalence<br>RS<sup>**</sup> (%) | Prevalence<br>SS<sup>***</sup> (%) | Quality<br>assessment |
| --- | --- | --- | --- | --- | --- | --- | --- | --- |
| Yoon KS<br>(19) | 2014 | USA | 291 | 90.72 | 85.98 | 14.02 | 10.23 | High |
| Durand R<br>(20) | 2011 | France | 167 | 93.41 | 98.08 | 1.92 | 7.05 | High |
| Kasai S<br>(21) | 2009 | Japan | 282 | 6.74 | 78.95 | 21.05 | - | High |
| Kwon DH<br>(22) | 2008 | Korea | 100 | 70.00 | 37.14 | 62.86 | 42.86 | High |
| Durand R<br>(23) | 2007 | France | 90 | 77.78 | 47.14 | 52.86 | 28.57 | Mild |
| Audino PG<br>(24) | 2005 | Argentina | 6250 | 15.20 | - | - | - | High |
| Kristensen<br>M (25) | 2005 | Denmark | 43 | 76.74 | 81.82 | 18.18 | 30.30 | Mild |
| Vassena<br>CV (26) | 2003 | Argentina | 26 | 92.31 | 85.98 | 14.02 | 10.23 | Mild |

Regarding the prevalence of kdr against organochlorine insecticides, 8 articles were included in the meta-analysis process, based on which, the prevalence of kdr was estimated at 65.3% (95% CI: 8.35-7.94). The highest prevalence of KDR was in the study conducted in France with a prevalence of 94% and the lowest prevalence was related to the study conducted in Argentina with a prevalence of 15.2%. which shows that the prevalence of kdr is different in different geographical areas (Figure 2).

**Figure 2:**
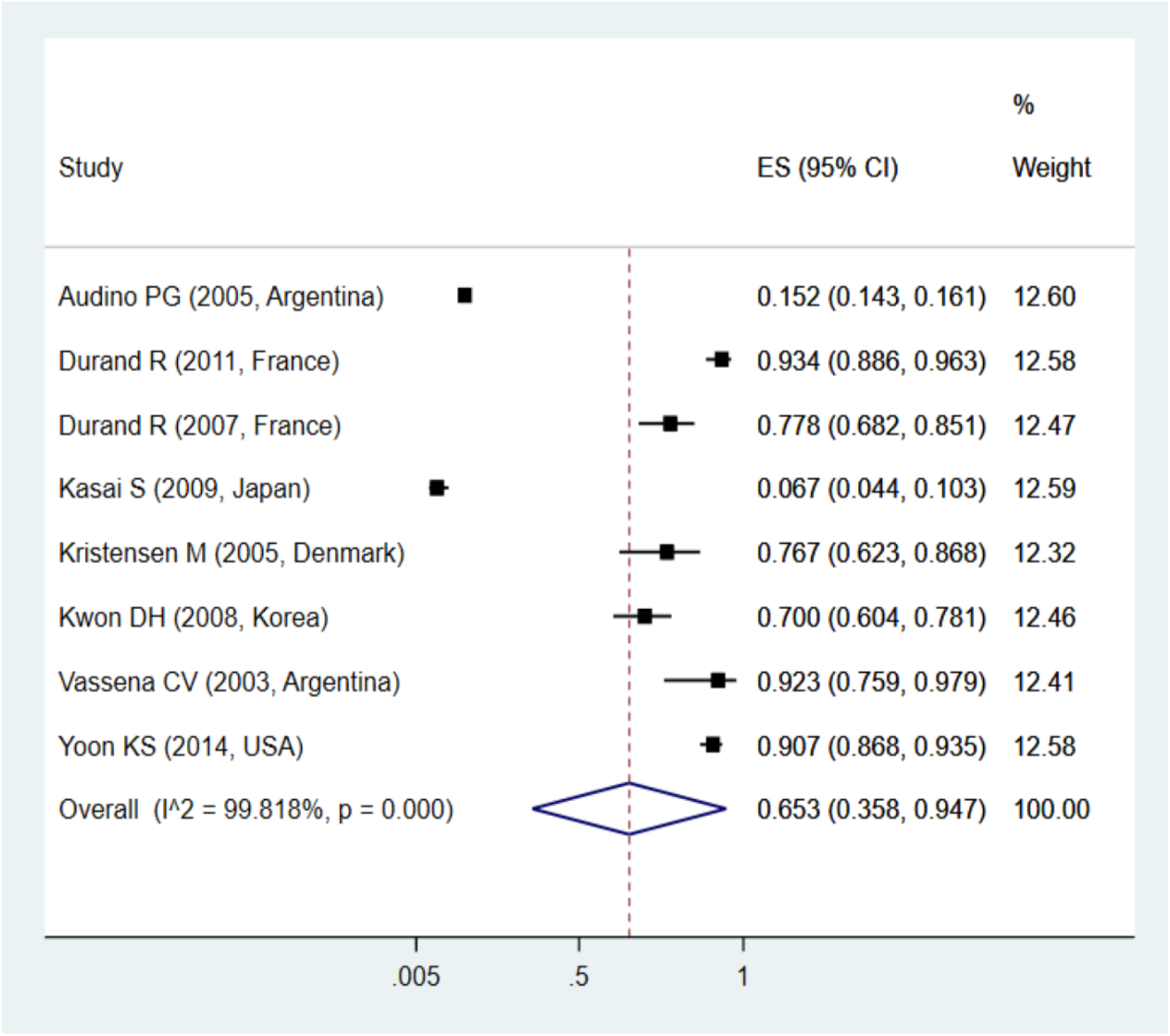
Pooled prevalence rate of Knockdown resistance based on random effects model. The midpoint of each line segment shows the prevalence estimate, the length of the line segment indicates the 95% confidence interval in each study, and the diamond mark illustrates the pooled prevalence of kdr.

The prevalence of kdr types by separating RR^1^, RS^2^, and SS^3^ alleles was studied in a meta-analysis. Based on the findings, the prevalence of RR was 71.9%, RS was 28.1%, and SS was 22.2%, which shows that most cases of resistance are of the RR type (Figures 3, 4, and 5).

**Figure 3:**
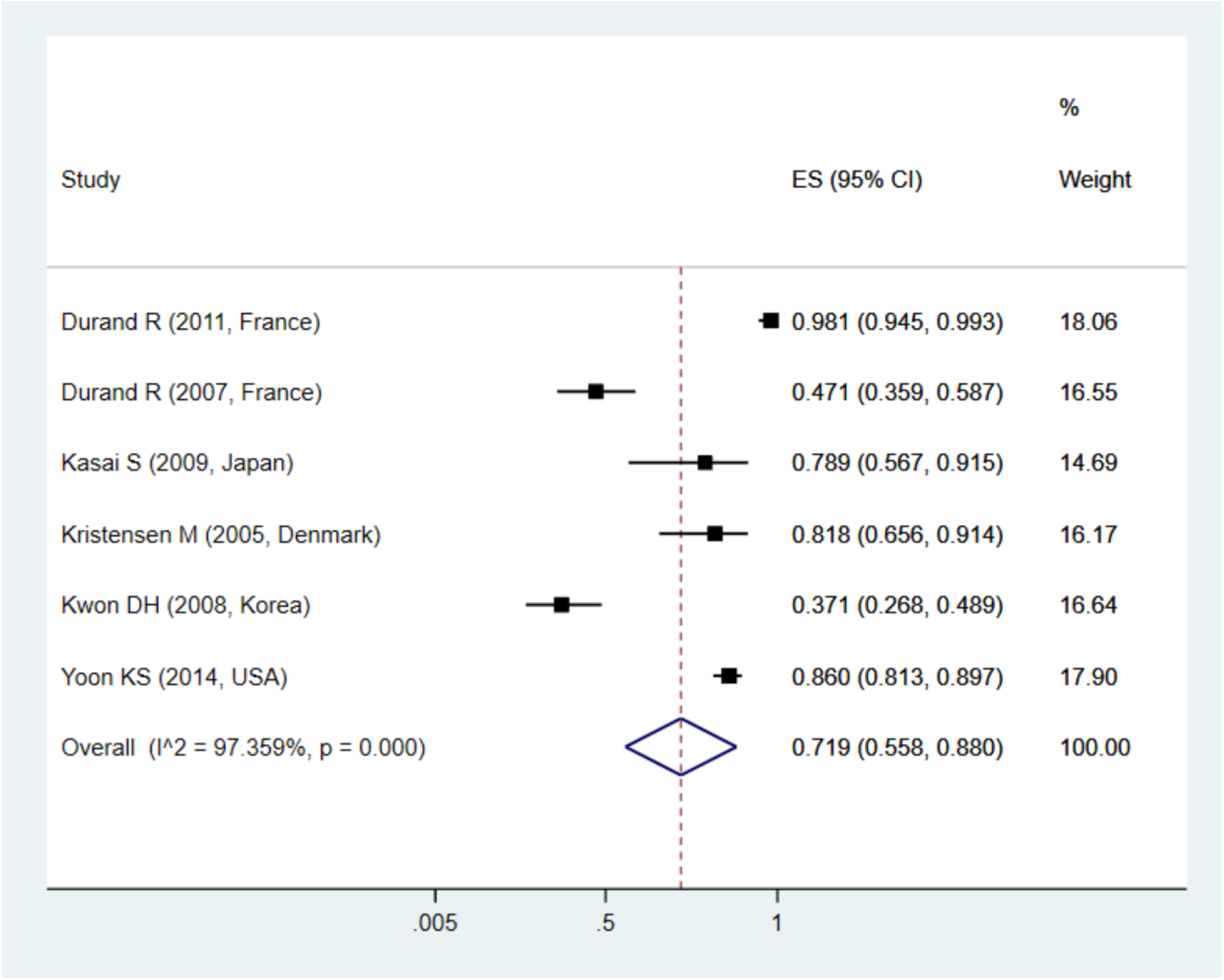
Pooled prevalence rate of RR based on random effects model. The midpoint of each line segment shows the prevalence estimate, the length of the line segment indicates the 95% confidence interval in each study, and the diamond mark illustrates the pooled prevalence of kdr.

**Figure 4:**
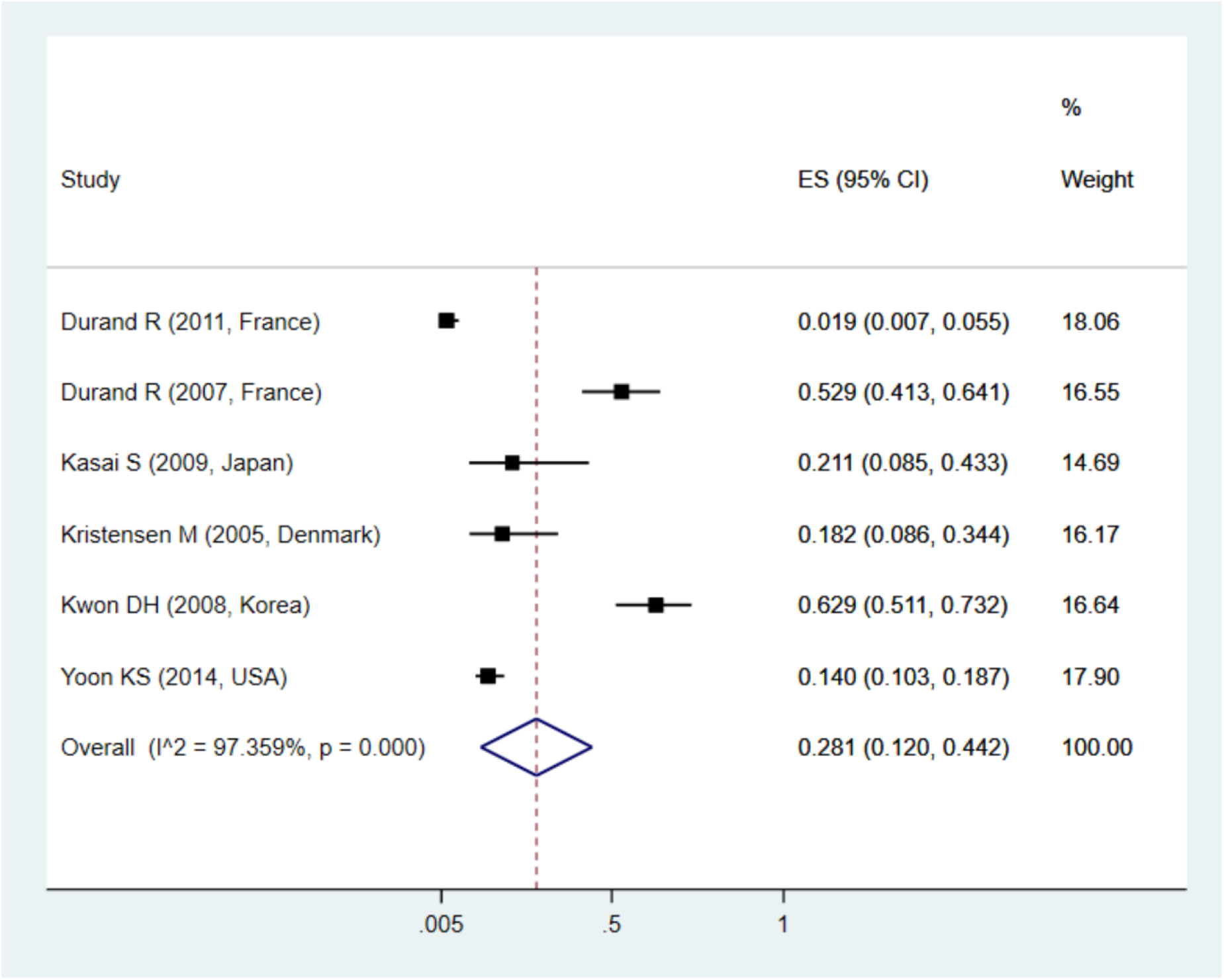
Pooled prevalence rate of RS based on random effects model. The midpoint of each line segment shows the prevalence estimate, the length of the line segment indicates the 95% confidence interval in each study, and the diamond mark illustrates the pooled prevalence of kdr

**Figure 5:**
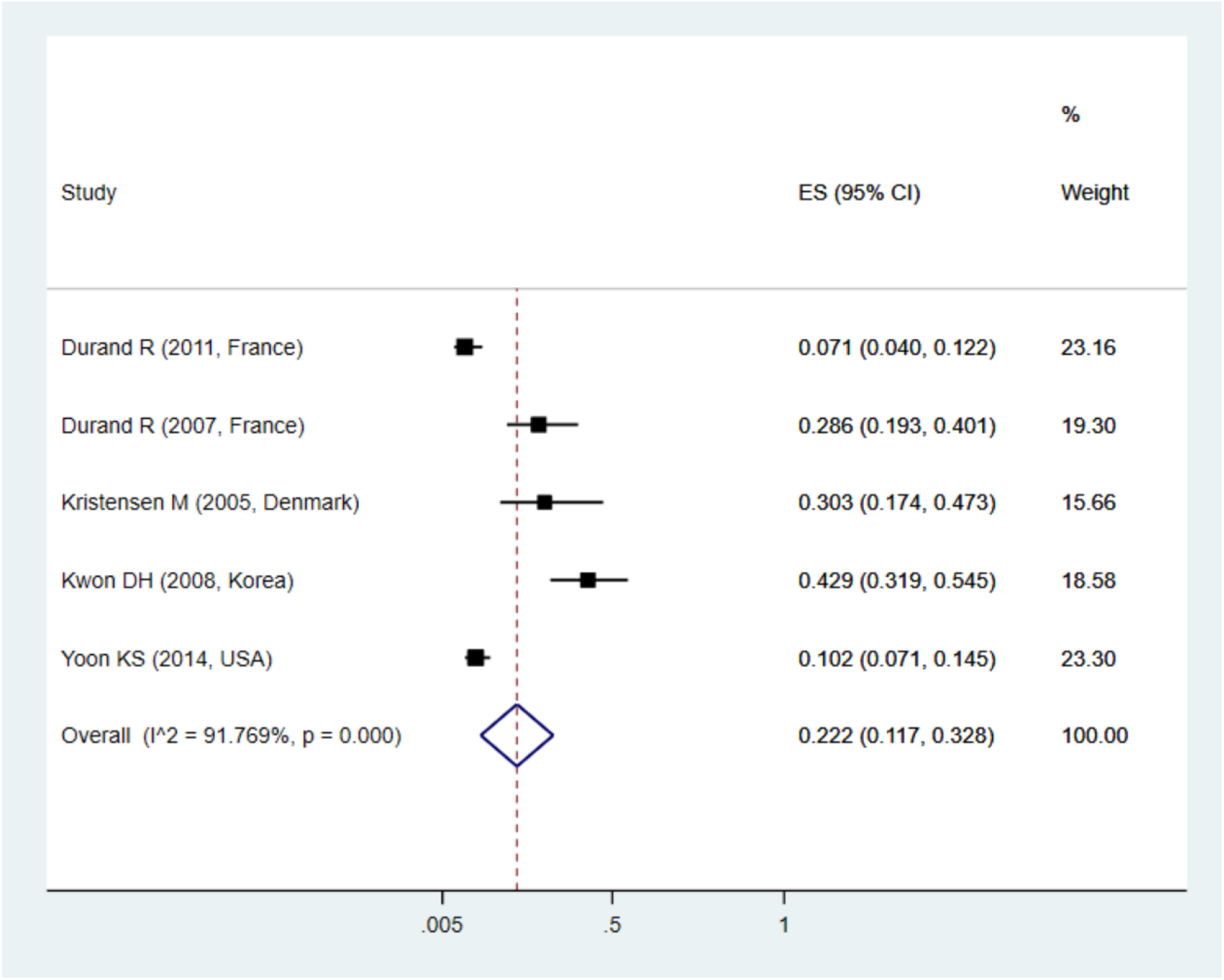
Pooled prevalence rate of SS based on random effects model. The midpoint of each line segment shows the prevalence estimate, the length of the line segment indicates the 95% confidence interval in each study, and the diamond mark illustrates the pooled prevalence of kdr.

**Figure 6:**
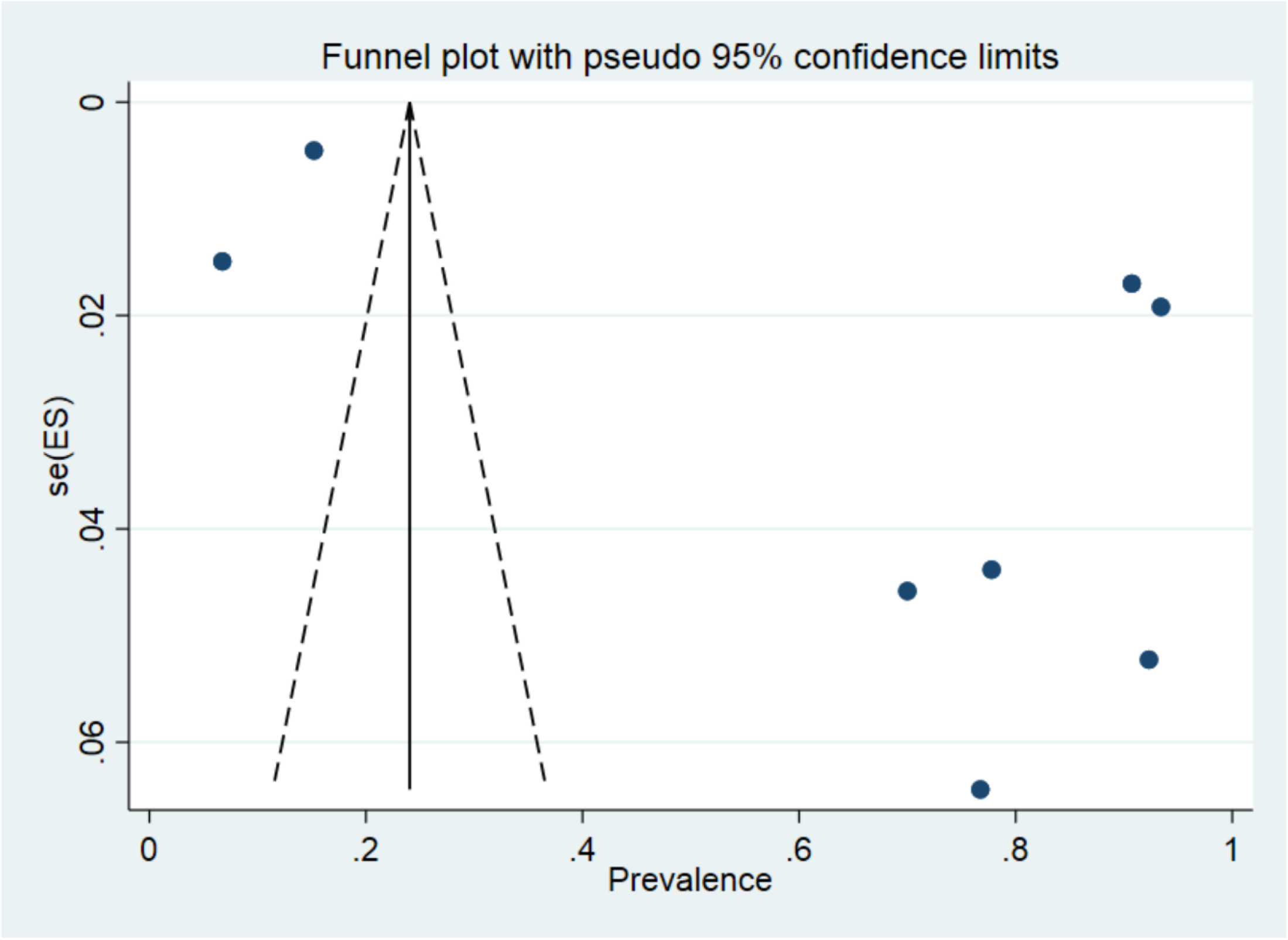
Funnel plot of the prevalence of kdr

Egger’s test and funnel plot were used to investigate the diffusion bias. Due to the diagram’s asymmetry, un needs to be published and accessed (P=0.01) (Chart 6). Meta-regression was used to investigate the relationship between the year of the study and the prevalence rate. According to the slope of the graph, the prevalence of kdr increased with the increase in the year of conducting the study, which shows that the prevalence of resistance to this group of insecticides is increasing (Figure7).

**Figure 7.**
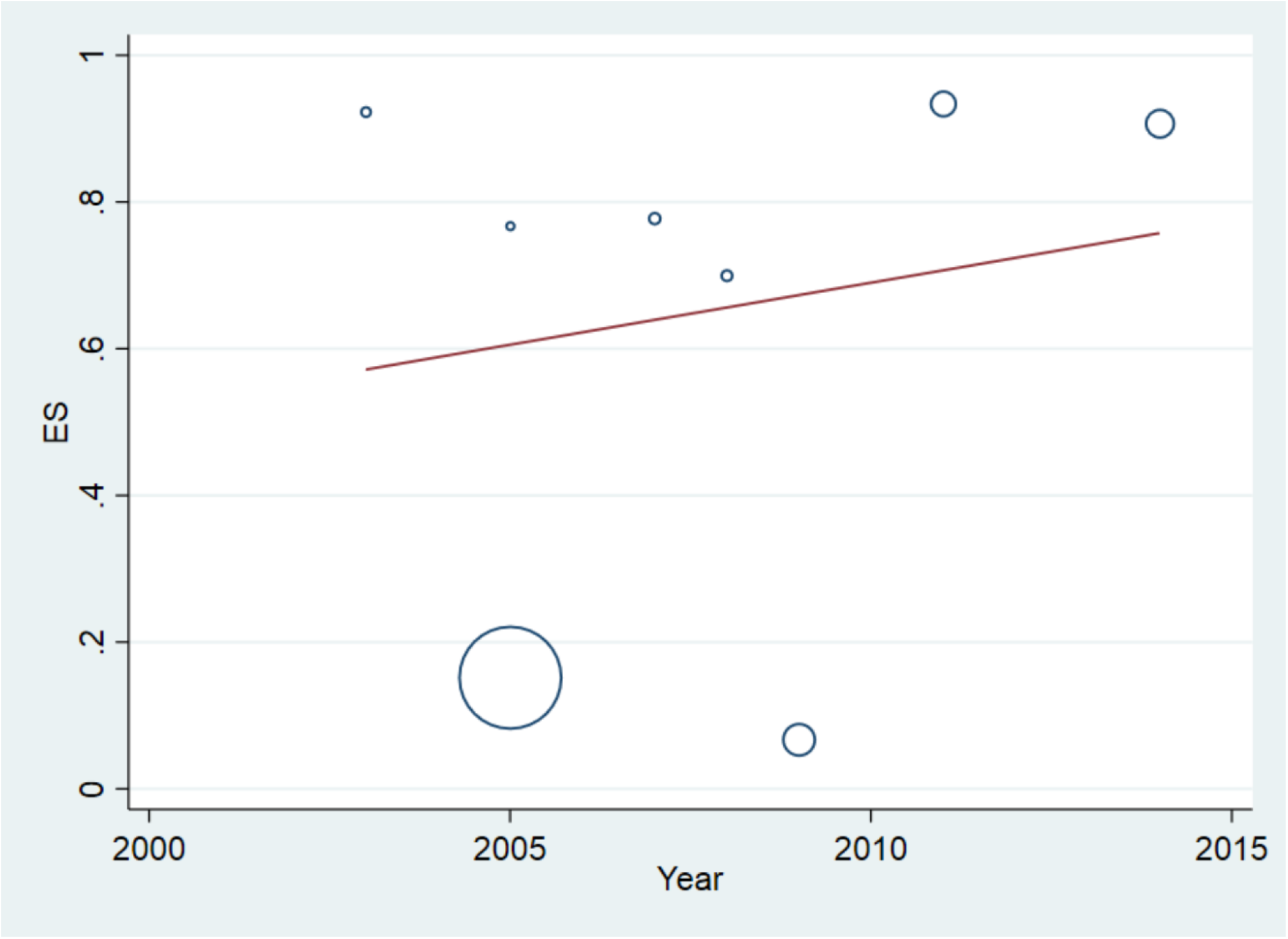
Meta-regression plot of the prevalence kdr based on the year of study

## Discussion

According to the findings of the present meta-analysis, about two-thirds of the human head lice population have kdr resistance against organochlorine insecticides. Based on this, the use of traditional and inappropriate treatment methods does not cause the treatment and control of this contamination but also leads to an increase in resistance against these insecticides. Although today, due to biological issues and resistance, organochlorine insecticides are not widely used to treat lice. But lindane was one of the insecticides that were used for treatment in the early years of the widespread outbreak of pediculosis, which is still used in some countries. But over the years, due to its widespread and unconventional use, the resistance to treatment with this insecticide increased so that in some studies the successful treatment with this insecticide reached less than 20% and its use in the European Union in It was banned in 2007. The cause of this was the lack of proper or incomplete treatment of this infection .(28, 27, 15) Another important issue is that when lice are under pressure, they can transfer resistance genes to each other. As a result, by transferring genes resistant to organochlorine insecticides to other lice, the issue of resistance to other insecticides such as pyrethroids is also raised, which is widely used for the treatment of pediculosis, and the mechanism of their effect is also through the effect It is on sodium channels. .(31-29)So resistance against this group of insecticides (pyrethroids) has also spread around the world .(15) .

Due to the existence of resistance to treatment in lice, today in the world, different methods are used to treat pediculosis. which include chemical treatment methods such as Permethrin 1%, Lindane 1%, Malathion 0.5%, (5)Physical methods such as occlusive agents and Dimethicone which lead to suffocation of the louse, Isopropyl myristate dissolves the surface wax of the louse’s body or desiccation which leads to the loss of water in the louse and its death, and manual removal which the louse uses The comb is separated from the hair (34-32)and the use of plant compounds such as Nopucid Bio Citrus, Nyda, Hedrin, and Nopucid Qubit, tea tree oil, eucalyptus oil, anethole, carvone, limonene, and linalool .(37-35)However, the use of treatment methods, especially chemical treatments, should be based on the principles and by completing the treatment period. Because if incomplete or single treatment is done and cannot eliminate the contamination, it can lead to the development of resistance develops. published studies Based on this, it is necessary to determine the resistance to insecticides and select the appropriate insecticide for the treatment before treating the infected areas. It is often recommended to use Polymerase chain reaction (PCR), Quantitative sequencing (QS), and real-time PCR (rtPASA) methods to determine resistance to treatment .(39, 38, 12) Generally, based on the present study and other conducted studies, the resistance to treatment in head lice against organochlorine insecticides is relatively high in the world, and their use is not recommended in some areas. As a result, using these insecticides alone in most cases can lead to treatment failure. Based on this, it is recommended that in different regions of the world, after determining the sensitivity of lice to insecticides, they should be used as a principle and combination of several insecticides for treatment and to prevent the spread of resistance in lice so that it can spread. reduced in the world.

## Conclusion

Meta-analysis findings showed that about 65% of human head lice are resistant to organochlorine insecticides. As a result, the presence of such high resistance affects the choice of these insecticides for treatment. Based on this, it is recommended to select the appropriate treatment, especially in the field of this group of insecticides.

## Data Availability

All data is included in the article.

## Footnotes

1 Homozygote resistant

2 Heterozygote resistant

3 Homozygous susceptible

